# The show must go on – what are the available tools to assess readiness to return to dance post injury in elite dancers? A scoping review protocol

**DOI:** 10.1101/2025.01.19.25320330

**Authors:** Róisín Cahalan, Ciarán Purcell, Rose Schmieg, Ms Edel Quin

## Abstract

Injury in elite dance is commonplace and pernicious. The decision to return to dance practice after injury requires careful consideration to ensure that the dancer is ready to safely resume practice. There are a limited number of pertinent assessment tools in dance, which largely consider predominantly physical domains and refer to specific dance genres only. Therefore, this scoping review aims to explore biopsychosocial domains identified in dance and sport literature to inform the safe return to practice post-injury in elite dance. The scoping review will conform to Joanna Briggs Institute (JBI) Evidence Synthesis guidelines. Ten databases will be searched for relevant articles. Two independent reviewers will conduct title and abstract screening followed by full-text screening. Data charting will be completing using a modified standardised form. Descriptive results will be reported using tabular and graphical media. The published scoping review will be disseminated to relevant stakeholders including those in the world of elite dance in both performance and educational settings, as well as to clinicians working with these artists. The resulting outputs will be in the form of both peer-reviewed and non-peer reviewed publications (e.g. blog posts, academic publications and conference presentations to reach key stakeholders such as dancers and their support teams). An infographic of key findings will be developed and shared on social media platforms as appropriate. The findings of this scoping review will inform a subsequent e-Delphi project involving dancers, teachers and clinicians to develop a dance-specific tool to inform safe return to dance post injury. Ethical approval was not required for this study.

- What is already known on this topic : Although injury in elite dancers is extremely common and often severe, there are limited tools available to guide safe return to dance after injury.
- What this study adds: By drawing on the broader literature in dance and sport, this study will identify key biopsychosocial domains that should be considered when deciding when a dancer is ready to safely resume dance practice after injury¡
- How this study might affect research, practice or policy: The outputs of this scoping review will raise stakeholder awareness of the biopsychosocial impacts of injury on dancers and will inform the development of a dance-specific instrument to help guide considered and appropriate return to dance post injury in this cohort.

## INTRODUCTION/BACKGROUND

Elite dancers are aesthetic athletes, coupling grace and intricate execution with a requirement for immense strength, power and cardiovascular fitness.^1^ As with many elite athletes, injury in dance is a constant and pernicious issue.^2^ The injury incidence rates reported in professional dancers has been found to range from 0.16 to 4.44 per 1000 hours of exposure with variation due to differing dance genres, injury definitions and load burden.^3-5^ The ability to accurately assess readiness for return to practice following injury is a critical aspect of injury management and rehabilitation in the dance community. Given the physical and technical demands placed on dancers, a comprehensive and reliable evaluation of readiness can aid in reducing the risk of re-injury, optimizing recovery outcomes, and ensuring safe reintegration into training and performance settings.

Despite the importance of this topic, there is a lack of consensus on which measures are most appropriate or effective in assessing dancers’ readiness to return to dance practice post-injury.^6^ The limited tools that are available in dance focus overwhelmingly on physical readiness to dance and overlook the many psychological and socioenvironmental issues that are inherent in the injury experience.^7^ Screening practices for injury in dance focus disproportionately on factors such as range of motion, flexibility, alignment and other physical attributes with minimal attention paid to psychosocial mediators of injury in dance cohorts.^8,9^ Regarding return to dance post-injury, possibly the most widely used tool in this space is the 14-item Dance Functional Outcome Survey (DFOS).^10^ This instrument interrogates a dancer’s ability to perform a host of general (e.g. walking, stairs, pain level) and technique (e.g. plié, développé, relevé balance etc.) elements, and has been established as a psychometrically sound tool to monitor lower extremity or low back injury in adult ballet and modern dancers. However, its utility in other dance genres or ability to establish the dancer’s psychological readiness to return to dance and impact of other contextual contributions to performance, is limited.

Research has indicated that numerous psychological factors including fear of return to practice post injury are useful determinants of successful functional return to dance.^11^ This reflects the research in sport where a systematic review of psychological factors associated with a return to sport post injury showed that positive psychological responses including motivation, confidence and low fear were associated with a greater likelihood of returning to the preinjury level of participation and returning to sport more quickly.^12^ Subsequent research identified that premature return to sport before the athlete was psychologically ready could result in a host of adverse outcomes including negative emotional states, suboptimal performance, increased re-injury risk, and long-term adverse career impacts.^13^ Thus, the importance of evaluating both the psychological as well as the physical status of the dancer or athlete cannot be overstated. There are numerous useful tools in sport including the Injury-Psychological Readiness to Return to Sport questionnaire^14^ and the Return to Sport After Serious Injury Questionnaire (RSSIQ)^15^ but validated tools in dance are lacking.

This scoping review therefore aims to systematically map and evaluate existing questionnaires, surveys and other relevant instruments that may be used to assess readiness to return to dance after injury, identify gaps in current methodologies, and highlight best practices in the field. By synthesizing the available literature, including pertinent tools for return to play after injury in sport (including performing arts athletes in circus/theatre/cheerleader settings), this review will provide an overview of physical, psychological, socioenvironmental and functional domains, addressing how these tools are used specifically within the dance context. Ultimately, this protocol will inform future research and contribute to the development of a standardized, evidence-based assessment framework tailored to the unique needs of dancers returning to practice after injury. An e-Delphi project based on the output of this scoping review involving dancers and teachers from multiple dance genres, as well as clinicians working in this area is planned. The aim of the e-Delphi exercise is to develop a dance-specific instrument to inform safe return to dance practice post injury. The instrument will be comprised of multiple domains that are common to all dance genres. A final domain that considers specific technique or choreographic elements applicable to individual dance genres will also be included allowing for tailoring to specific needs of dancers from different styles.

## METHODS AND ANALYSIS

This scoping review will be conducted in accordance with the Joanna Briggs Institute (JBI) Evidence Synthesis guidelines^16^ and Preferred Reporting Items for Systematic Reviews and Meta-Analyses extension for Scoping Reviews (PRISMA-SCr).^17^ Building on a prior framework and methodological guidance, this approach facilitates enhanced development and reporting of appropriate objectives and comprehensive search strategies. Precision of the topic was further determined through the use of an expanded population, concept, instrument and context paradigm to focus the title, aims and objectives of the review. This facilitated a comprehensive search strategy, enhanced transparency and rigour of reporting and synthesis and presentation of findings (Table 1).

**Table 1:** Key concepts informing search strategy.

| Population | Concept (Outcome/ Condition) | Context | Instrument/ Tool |
| --- | --- | --- | --- |
| Dancers (and synonyms) | Readiness to return to sport/dance | Dance injury | Questionnaire, survey, scale, tool, inventory, assessment, measure, instrument |
| Athletes (and synonyms) | Return-to-play (RTP) / Return-to-dance (RTD) | Sport injury |  |
| Performance athletes (e.g. circus/theatre athletes, cheerleaders). | Rehabilitation and recovery | Musculoskeletal injury |  |
|  | Physical readiness, psychological readiness | Rehabilitation and recovery |  |
|  | Outcome measures for readiness |  |  |

### Inclusion and exclusion criteria

The following criteria will be applied to the search in this scoping review:

### Inclusion criteria

- Original peer-reviewed research, systematic reviews, and clinical practice guidelines reporting assessment or evaluation tools (including surveys, questionnaires) of readiness to return to performance post injury.
- Studies including sport, dance or other physical performance (e.g. circus/theatre/cheerleading).
- English language, human research studies published from the date of inception of the database.
- Elements of return to performance tools for specific injuries (e.g. ACL) pertaining to non-specific physical elements of the injury will be included. For example, psychological and general functional readiness may be included.

### Exclusion criteria

- Studies focussing solely on concussion will be excluded, due to the low occurrence of concussion in dance.
- Review articles, conference proceedings and other secondary sources will be excluded.

These criteria have been applied to identify a breath of tools which encompass numerous holistic domains and may be used by non-clinicians as well as experienced professionals, to inform decision making regarding return to dance. By including condition-specific return to practice tools, we hope to identify domains that can be more widely applied to broader injury diagnoses in dancers.

#### Information Sources and Search Strategy

The search strategy has been developed by the research team, who share a wealth of clinical and research experience in dance and sport. The search strategy aims to locate published primary studies alongside systematic reviews and clinical practice guidelines. An initial limited search of Web of Science and CINAHL Ultimate (EBSCO) was undertaken to identify articles on the topic. The text words contained in the titles and abstracts of relevant articles, and the index terms used to describe the articles were used to develop a full search strategy for MEDLINE (EBSCO) (Appendix I, supplementary material).

The search strategy, including all identified keywords and index terms, will be adapted for each database from inception; Databases are selected based on their relevance to the research topics of health, medicine, kinesiology, sport and dance. These databases include: Web of Science; EMBASE; Cochrane Database of Systematic Reviews; EBSCO (CINAHL Ultimate, MEDLINE, SPORTDiscus, PsycINFO); PubMed; Elsevier (ScienceDirect); ProQuest Performing Arts Periodical Database, Dissertations); SAGE; JSTOR; and PEDro: the Physiotherapy Evidence Database. Grey literature will also be searched for pertinent articles (OpenGrey and GreyLit.org).

### Study Selection and Screening

The reference lists of articles included in the review will be screened for additional papers. Studies will be limited to those published in English and on human subjects. Following the search, all identified records will be collated and uploaded into Endnote X9.3.3(Clarivate Analytics, PA, USA) and duplicates will be removed. Following a pilot test, titles and abstracts will then be screened by two independent reviewers (RC & CP) for assessment against the inclusion and exclusion criteria for the review. Potentially relevant papers will be retrieved in full and their citation details imported into the Covidence reference management system. (Covidence; Covidence Melbourne, Australia). The full text of selected citations will be assessed in detail against the inclusion and exclusion criteria by two independent reviewers (RC & CP). Reasons for exclusion of full-text papers will be recorded and reported in the scoping review. Any disagreements that arise between the reviewers at each stage of the selection process will be resolved through discussion or with a third reviewer (RS). Authors of papers will be contacted to request missing or additional data, where required. If access to missing data is not possible these papers will be excluded. The results of the search will be reported in full in the final scoping review and presented in a Preferred Reporting Items for Systematic Reviews and Meta-analyses for Scoping Reviews (PRISMA-ScR) flow diagram.^17^

### Data Charting/Collection/Extraction

Data charting, collection, and extraction for this scoping review will follow a systematic and transparent process to ensure comprehensive capture of relevant information.^16^ A standardized data extraction form will be developed, detailing key information such as the type of tool (e.g., questionnaire, assessment protocol), the target population (e.g., dancers, athletes), domain examined (e.g., function, symptoms, psychological factors); the specific injury or condition addressed, the purpose of the tool (e.g., assessing readiness to return to activity, guiding rehabilitation). Data will be extracted from eligible studies by two independent reviewers (EQ & RS) to minimize bias, with discrepancies resolved through discussion or consultation with a third reviewer. Data will be charted in a systematic matrix or table for easy comparison, summarizing key domains to inform the development of a future tool for return to dance. This matrix will be modified and revised as necessary during the data charting process with any modifications highlighted in the full scoping review.

### Synthesis and Presentation of Results

A PRISMA flow diagram will document the selection process, from database searching and article screening to the final selection of studies for inclusion in the review. Selected data will be extracted and presented in a table with fields including study details (author/year), population, nature of the tool, nature of injury, domains included, purpose of the tool, administration details, and any associated psychometric properties where available (Appendix 2, Supplementary material). Data synthesis will involve categorizing and summarizing the key findings of the included studies, such as the types of instruments used, domain properties, populations studied, and any identified gaps or challenges in establishing readiness for return to practice. Descriptive analysis with basic coding will be used to present the type and frequency of common domains used across studies, which will be presented in table or graphical format.

## Discussion and Dissemination

Injury in dance is extremely common, but there is a lack of guidance for stakeholders to inform the decision to return to dance after injury. Existing tools in dance focus largely on physical readiness to return to dance, eschewing important psychosocial considerations. Additionally, genres apart from ballet are poorly addressed. This scoping review will explore existing instruments in dance and sport to identify a range of domains that should be considered when assessing the dancer’s holistic preparedness to resume dance practice. In doing so, we aim to raise stakeholder awareness of the complexity of factors that inform return to dance decisions. As mentioned, outputs will further inform the development of a dance-specific instrument to guide these decisions.

The results of this scoping review will be disseminated in both academic and dance fora. An academic paper will be submitted for peer reviewed publication, and to appropriate conference(s). A lay summary will also be made available to a non-academic audience and distributed to the professional and dance education communities. These include dance and performing arts conservatoires, companies and higher education institutions. Once published the results of the study will be summarised and shared in plain language to dancers in digital format (X, Instagram) and through the Universities of the authors and affiliated dance organisations.

## Data Availability

This is a study protocol and therefore no data has been generated apart from search strategies which are available in the present work.

