## Supplemental material for "The show must go on – what are the available tools to assess readiness to return to dance post injury in elite dancers? A scoping review protocol"

**Supplementary material**

**Appendix 1: Key Terms and Synonyms**

**a) Population (athletes and dancers):**

- **Dancers**: (MeSH: "Dancers") OR dancer* OR "ballet" OR "professional dancers"
- **Athletes**: (MeSH: "Athletes") OR athlete* OR "sports" OR "sport*" OR "competitive athlete*"

**b) Readiness to return (to sport or dance):**

- **Readiness**: (MeSH: "Psychological Readiness") OR "return to sport" OR "return to dance" OR "return to play" OR "sport participation" OR "post-injury rehabilitation" OR "physical readiness" OR "psychological readiness"

**c) Instrument/Measurement tool:**

- **Measurement tools**: (MeSH: "Questionnaires") OR "survey" OR "questionnaire" OR "scale" OR "inventory" OR "tool" OR "instrument" OR "assessment" OR "measure" OR "evaluation"

**d) Injury context:**

- **Injury context**: (MeSH: "Sports Injuries") OR "musculoskeletal injury" OR "rehabilitation" OR "injury recovery" OR "physical therapy" OR “dance injury”

<u>**Search Strings**</u>

(

(ti("athlete*") OR ab("athlete*") OR tw("athlete*") OR ti("dancer*") OR ab("dancer*") OR tw("dancer*"))

**AND**

(

ti("readiness to return") OR ab("readiness to return") OR tw("readiness to return")

OR ti("return to play") OR ab("return to play") OR tw("return to play")

OR ti("return to dance") OR ab("return to dance") OR tw("return to dance")

OR ti("psychological readiness") OR ab("psychological readiness") OR tw("psychological readiness")

OR ti("physical readiness") OR ab("physical readiness") OR tw("physical readiness")

)

**AND**

(

ti("questionnaire") OR ab("questionnaire") OR tw("questionnaire")

OR ti("survey") OR ab("survey") OR tw("survey")

OR ti("scale") OR ab("scale") OR tw("scale")

OR ti("inventory") OR ab("inventory") OR tw("inventory")

OR ti("assessment") OR ab("assessment") OR tw("assessment")

OR ti("measure") OR ab("measure") OR tw("measure")

OR ti("tool") OR ab("tool") OR tw("tool")

OR ti("instrument") OR ab("instrument") OR tw("instrument")

)

**AND**

(

ti("sports injury") OR ab("sports injury") OR tw("sports injury")

OR ti("musculoskeletal injury") OR ab("musculoskeletal injury") OR tw("musculoskeletal injury")

OR ti("rehabilitation") OR ab("rehabilitation") OR tw("rehabilitation")

OR ti("injury recovery") OR ab("injury recovery") OR tw("injury recovery")

)

)

### Appendix 2: Presentation of extracted data

| Study Details | Population | Nature of Tool | Injury/ Condition | Domains | Purpose of Tool | Administration details | Psychometric Properties |
|---|---|---|---|---|---|---|---|
